# Temporal Changes in Immunization Information Systems Across U.S. States and Jurisdictions, 2000-2024

**DOI:** 10.64898/2026.05.29.26354476

**Authors:** Tianci Chen, Miwa Watanabe, Timothy Callaghan, Kayoko Shioda

## Abstract

**Background:** Statewide immunization data are essential for monitoring vaccination trends and evaluating immunization program impact. In the United States, Immunization Information Systems (IIS) were established in the early 1990s to collect these data; however, operational, legal, and procedural details vary across states and over time. This study summarized differences in IIS characteristics, such as legal requirements and reporting procedures, across U.S. states and jurisdictions over time.

**Methods:** We analyzed survey data from previous work in 2000 and the Centers for Disease Control and Prevention (CDC) in 2012, 2018, and 2024. Our review focused on legislation and reporting requirements for immunization registries across 50 states and 14 jurisdictions, including U.S. territories and Freely Associated States.

**Results:** Between 2000 and 2024, legal frameworks and reporting practices for immunization registries expanded across U.S. states and jurisdictions. The number of states with laws or administrative rules authorizing immunization registries increased from 24 states in 2000 to all 50 states, the District of Columbia, five metropolitan areas, five U.S. territories, and three Freely Associated States in 2024. Over the same period, reporting requirements also became more widespread. The number of states and jurisdictions mandating providers to report immunization records increased from 12 in 2000 to 54 in 2024. Consent policies also changed over time. By 2024, most states and jurisdictions had adopted implicit consent for reporting children’s immunization records (41; 64%), while a smaller proportion required explicit parental consent (7; 11%) or implemented mandatory reporting without consent (14; 22%).

**Discussion:** IIS infrastructure and reporting requirements have expanded across U.S. states and jurisdictions over the past two decades, while heterogeneity in consent policies and reporting practices persists. These temporal changes may need to be considered when interpreting IIS data, particularly in longitudinal and cross-jurisdictional analyses.

## Introduction

National and regional immunization data are essential for monitoring vaccination trends, evaluating the impact of vaccination programs, and informing public health decision-making. In the United States, efforts to develop immunization registries in states and local communities began in the early 1990s [1–3]. Today, states and jurisdictions use Immunization Information Systems (IIS), which are secure, population-based electronic systems to collect and manage data on vaccinations administered by participating providers in a specific jurisdiction [4]. IIS is useful at both the individual and population levels. At the individual level, IIS compiles vaccination histories from multiple sources, which providers can use to determine appropriate vaccinations for their patients. At the population level, aggregated IIS data can be used for surveillance and program operations to guide public health actions.

IIS provides important data for vaccine evaluation. For example, recent studies have used IIS data to assess the safety and efficacy of COVID-19 vaccines in 18 U.S. states participating in the IVY Network [6,7] and rotavirus vaccines in Connecticut [8]. However, to conduct national and longitudinal vaccine evaluation, it is important to understand regional and temporal differences in IIS regulations and data collection methods, as these differences may affect the comparability of IIS data [9]. Therefore, this study reviewed and summarized how the regulations and policies governing IIS changed across jurisdictions and over time in the years 2000, 2012, 2018, and 2024.

## Methods

### Data sources

We reviewed data on state and jurisdictional regulations related to IIS for the years 2000, 2012, 2018, and 2024.

Data for the year 2000 were extracted from Horlick *et al*. [1], which compiled information through telephone interviews conducted in 1997 and 1998 with immunization program administrators or their designees at state health departments, followed by a data update in 2000. The study also incorporated information from legislative documents, administrative regulations, and immunization policy records.

Data for 2012 and 2018 were extracted from Martin et al. and the CDC [9,11], which were collected through an online survey and follow-up telephone interviews conducted by the Institute for Public Health Informatics on behalf of the CDC National Center for Immunization and Respiratory Diseases and IIS Support Branch. These surveys were administered to immunization program staff to gather information on legislation governing IIS operations.

Data for 2024 were extracted from the CDC website [5], which contained data from a survey conducted by the CDC’s Informatics and Data Analytics Branch. This survey, distributed via email to state and jurisdictional immunization programs, collected information on consent procedures, provider reporting requirements, and data-sharing policies related to IIS.

In 2000, data were collected from all 50 states and Washington, D.C (n=51 jurisdictions). In 2012, all 50 states as well as Washington, D.C., Philadelphia, and San Antonio, were included (n=53 jurisdictions). In 2018, 49 states (other than New Hampshire) as well as Washington, D.C., Guam, New York City, Philadelphia, and San Antonio were included (n=54 jurisdictions). In 2024, 50 states as well as Washington D.C., New York City, Philadelphia, San Antonio, Houston, and five U.S. territories (American Samoa, Guam, the Commonwealth of the Northern Mariana Islands, the Commonwealth of Puerto Rico, and the U.S. Virgin Islands), District of Columbia, and three freely associated states (the Federated States of Micronesia, the Republic of the Marshall Islands, and the Republic of Palau) were included (n=64 jurisdictions).

It should be noted that the immunization registry data collected in the previous study [1] focused on state immunization registries rather than specifically on IIS. Therefore, some states may have operated their own immunization registries in 2000 without participating in the IIS network monitored by CDC. For example, New Hampshire had legislation authorizing an immunization registry in 2000, but it did not participate in IIS until later. We still included such jurisdictions in the analysis to capture temporal differences in immunization registry policies across jurisdictions. In contrast, the data collected for 2012-2024 specifically focused on IIS across jurisdictions; therefore, jurisdictions that did not participate in IIS during those years were not included (e.g., New Hampshire in 2012 and 2018).

### Extracted information

Information on key characteristics of IIS regulations for each state and jurisdiction in each study year was extracted from the data sources. First, we collected data on age groups authorized for inclusion in immunization records (children only vs. all ages). Second, we summarized reporting mandates for providers, including public health providers, Vaccines for Children (VFC) providers, private providers, and pharmacists/pharmacies, as well as the entities to which they were required to report (IIS or local public health authorities). Third, we reviewed consent procedures for child and adult immunization recipients, classifying them as no consent, implicit consent, or explicit consent. We also extracted details on the type of explicit consent (verbal, written, or either) in 2000, 2012, and 2018, while this information was not available for 2024. Finally, we recorded whether states and jurisdictions had regulations governing data sharing.

### Temporal trend analysis

For states and jurisdictions with at least two data points between 2000 and 2024, we evaluated how key IIS characteristics changed over time. Specifically, we assessed whether each state or jurisdiction had no change, increased/expanded, or reduced/narrowed reporting requirements in five areas: age group coverage, provider reporting mandates, consent for children, consent for adults, and data sharing policies. An “increased/expanded” policy trend refers to expanded population enrollment in state IIS; shifts in regulations from IIS participation with explicit consent to implicit consent to mandatory participation without consent; and the introduction of data sharing regulations. A “reduced/narrowed” policy trend refers to narrowed population enrollment in state IIS; shifts in regulations from mandatory participation without consent to participation with implicit consent to explicit consent; and the removal of data sharing regulations.

## Results

### Age groups covered

With respect to age groups, most states and jurisdictions were authorized to collect immunization records for all age groups in all years (**Table 1**). From 2000 to 2018, Connecticut limited inclusion to children under 6 years old, and Rhode Island limited inclusion to residents under 19 years old, but both expanded to all age groups by 2024 (**Figure 1-A**).

**Table 1.**
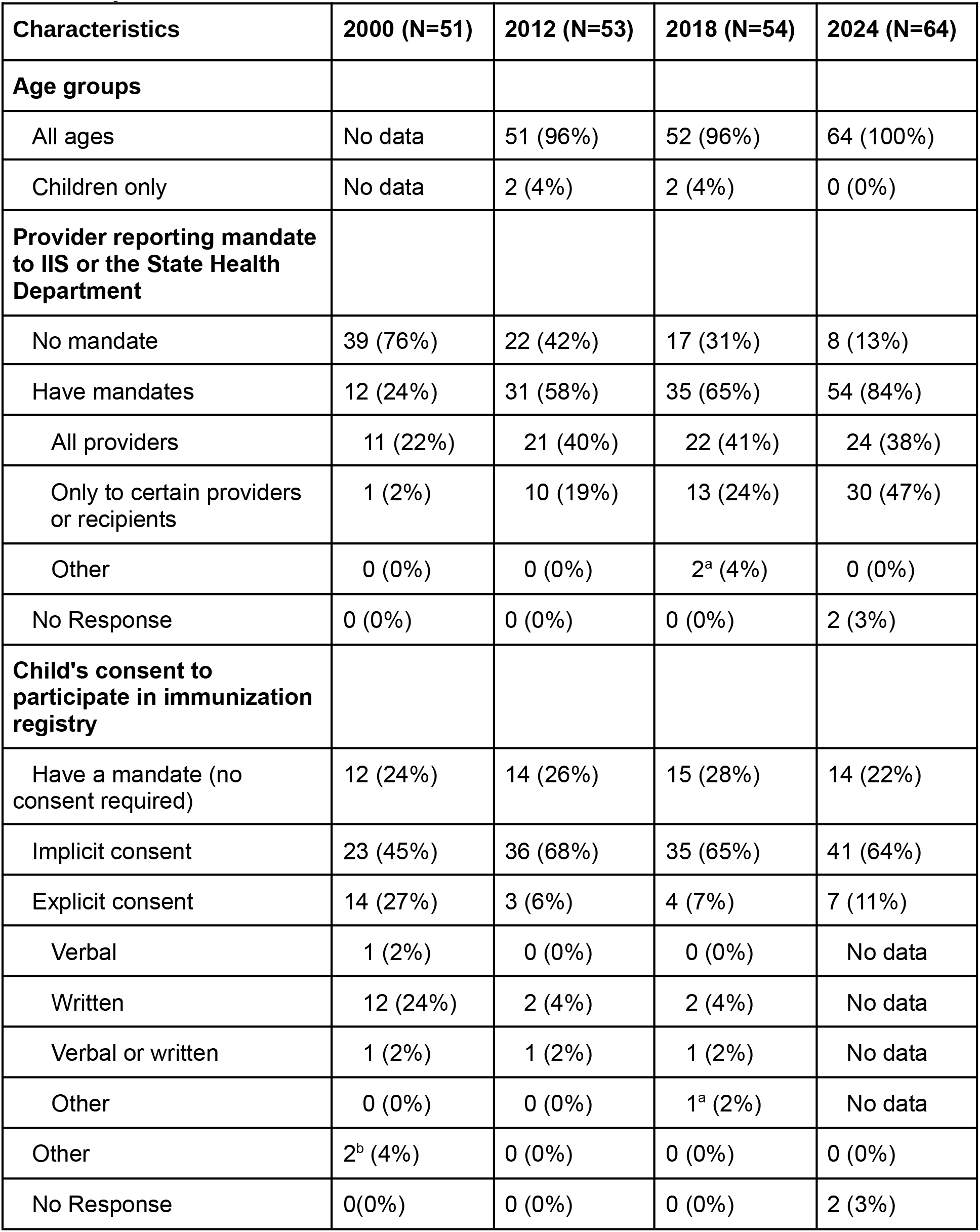
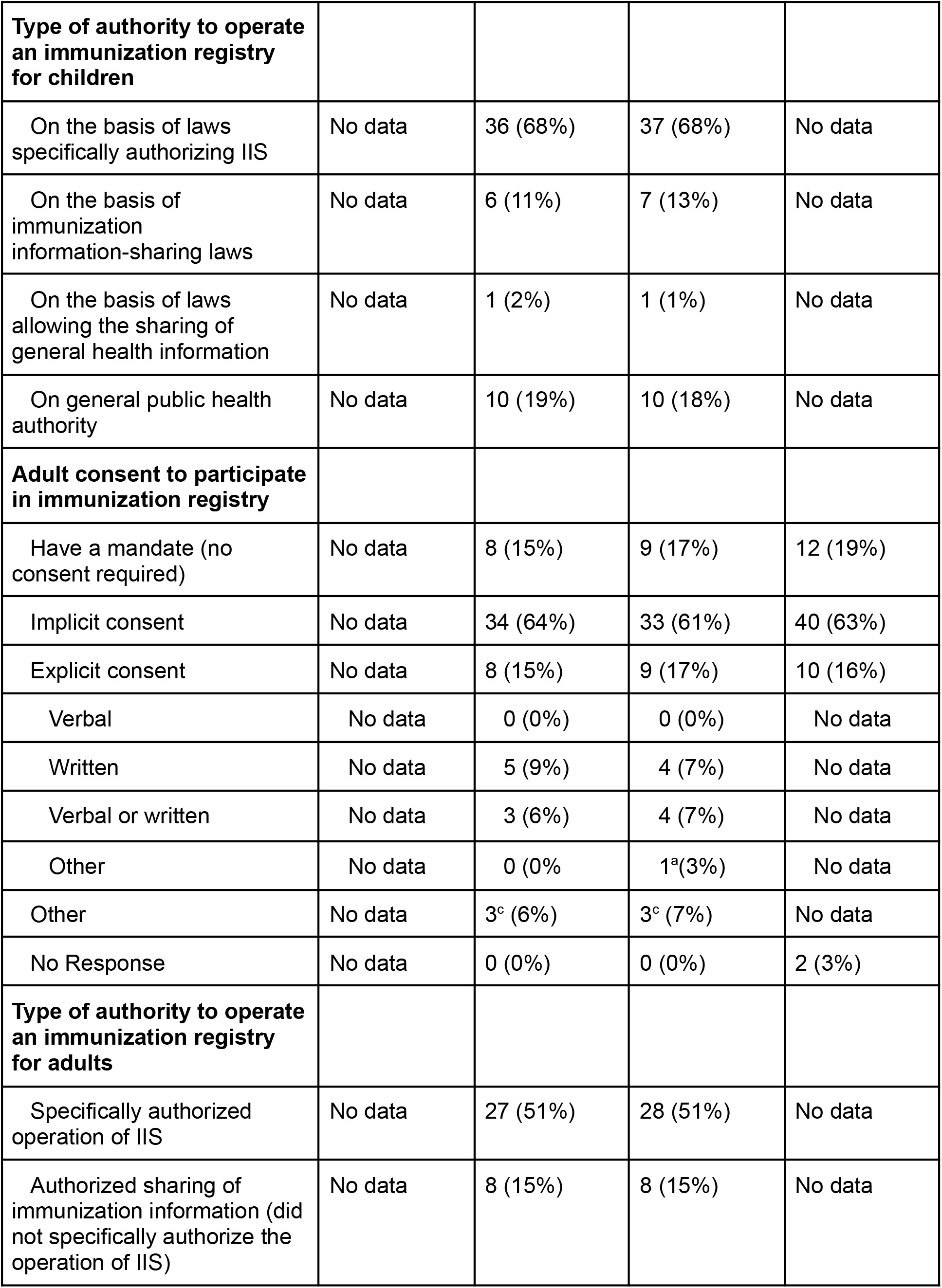
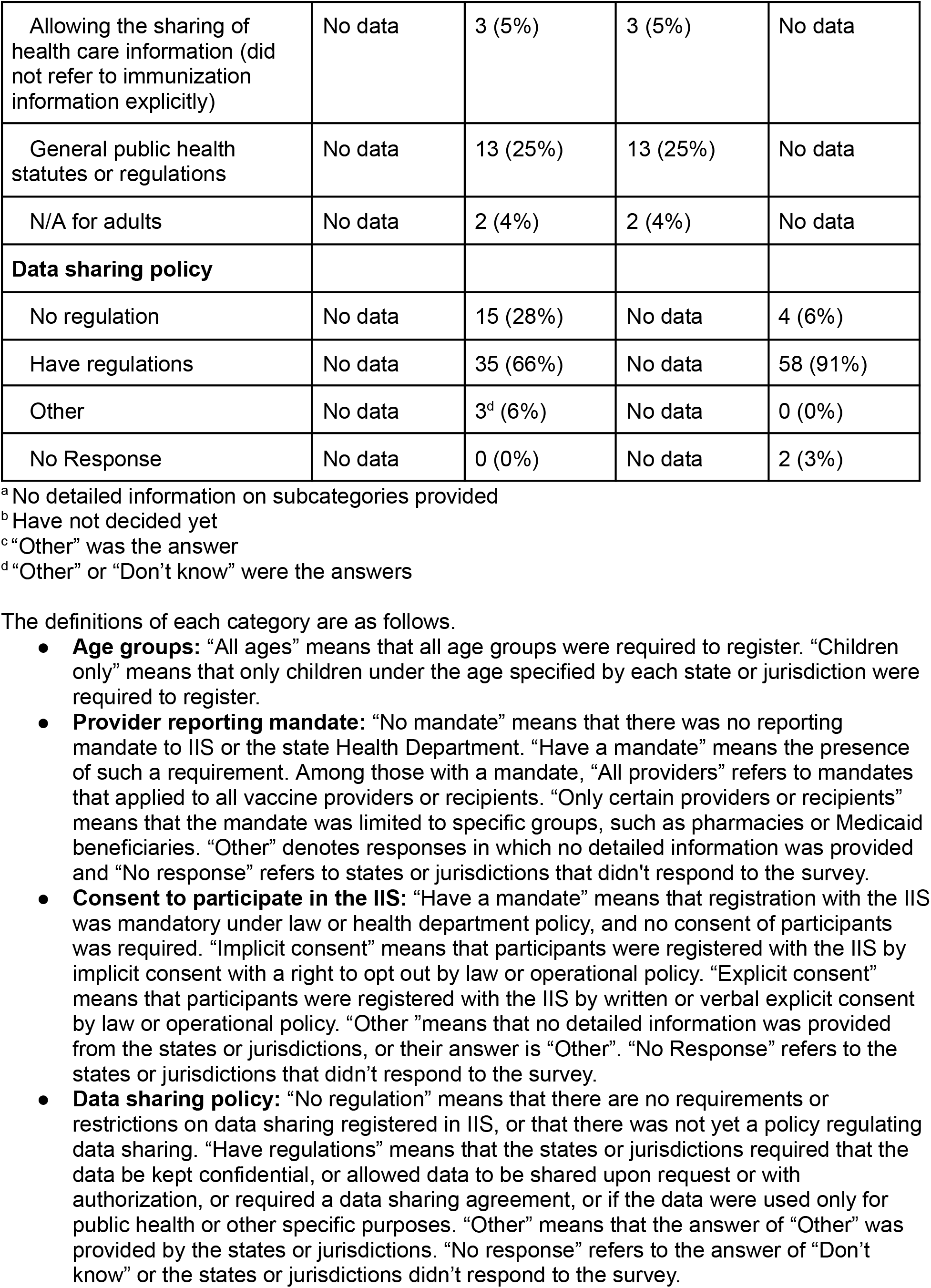
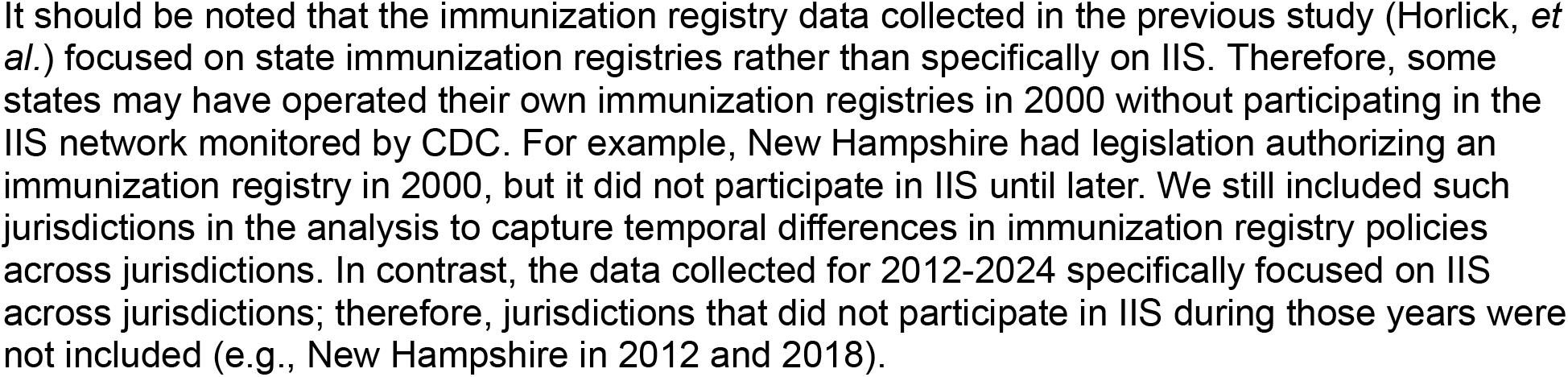
Key characteristics of regulations regarding Immunization Information Systems across states and jurisdictions in 2000, 2012, 2018, and 2024.

**Figure 1.**
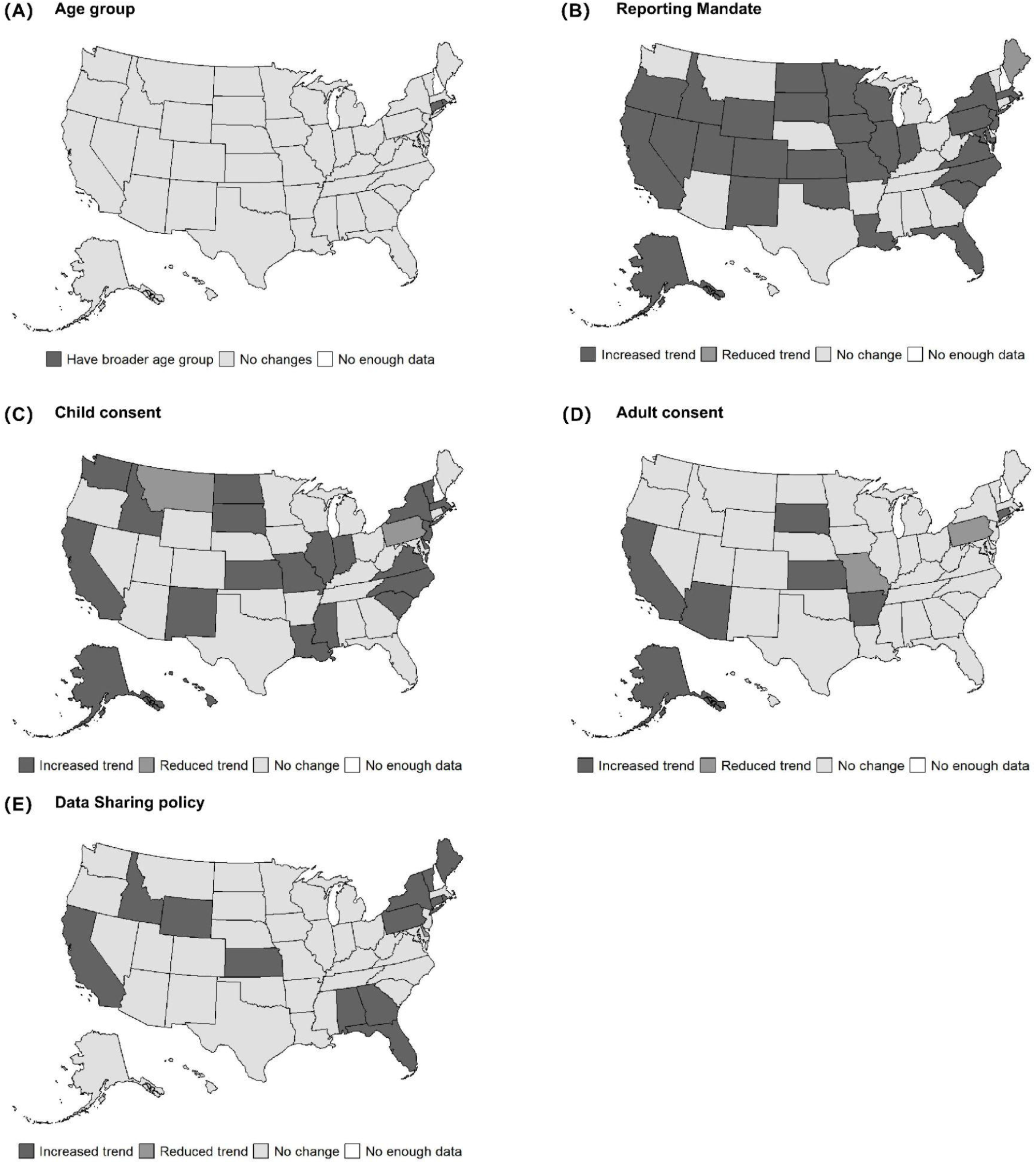
Trends in Immunization registries or IIS regulation changes among states that had more than two data points between 2000 and 2024.

### Immunization reporting mandates

Reporting mandates for immunization records varied across states and jurisdictions, with an overall increase in reporting mandates over time (**Table 1, Figure 1-B**). The majority of states and jurisdictions required at least one type of provider or entity to report immunizations, increasing from 12 (24%) in 2000 to 54 (84%) in 2024.

With respect to the types of reporters, more than half of states and jurisdictions (n=54, 84%) required reporting from certain providers in 2024. The number of states and jurisdictions requiring all immunization providers to report to IIS increased over time, from 11 states and jurisdictions (22%) in 2000 to 24 (38%) in 2024. Similarly, the number of states and jurisdictions requiring mandatory reporting from only certain providers or recipients increased from 1 (2%) in 2000 to 30 (47%) in 2024.

### Consent to share childhood immunization information

Between 2000 and 2024, the number of states and jurisdictions requiring consent to share childhood immunization information increased from 39 (76%) to 50 (78%), while the remaining jurisdictions allowed childhood immunization information to be shared with the IIS without consent (**Table 1, Supplementary Figure 1**). Regarding consent models, the proportion requiring implicit consent with opt-out increased over time, from 23 (45%) in 2000 to 41 (64%) in 2024. In contrast, the number of states requiring explicit consent with opt-in decreased from 14 (27%) in 2000 to 3 (6%) in 2012, before increasing to 7 (11%) by 2024. Of those requiring explicit consent, 12 (24%) required written consent in 2000, which became less common in 2012 (n=2, 4%) and 2018 (n=2, 4%).

Consent models changed over time in many states, with shifts in both directions (**Figure 1-C**). Some states moved toward more comprehensive enrollment policies. For example, Alaska, California, and South Dakota transitioned from implicit consent with opt-out in 2012 to mandatory participation without consent by 2018 or 2024. Kansas shifted from explicit written consent in 2012 and 2018 to implicit consent with opt-out in 2024. In contrast, other states moved toward more restrictive consent models. Missouri, Oregon, and Rhode Island required mandatory participation in 2012 and 2018 but transitioned to implicit consent with opt-out in 2024. Pennsylvania shifted from implicit consent with opt-out in 2012 and 2018 to explicit consent with opt-in in 2024. New Hampshire consistently required explicit consent with opt-in for children in 2018 and 2024.

### Consent to share adult immunization information

Similar to child immunization information, consent for the registration of immunization data was required for adults in about 80% of states and jurisdictions in each year (**Table 1, Supplementary Figure 2**). The number requiring implicit consent increased from 34 (64%) in 2012 to 40 (63%) in 2024, and explicit consent increased from 8 (15%) in 2012 to 10 (16%) in 2024.

Although the overall number of states requiring implicit or explicit consent remained stable, several states shifted between implicit and explicit consent models over time (**Figure 1-D**). For example, Arizona, Arkansas, and Kansas required explicit written consent in 2012 and 2018, but in 2024, they required implicit consent. Pennsylvania required implicit consent in 2012 and 2018, but it required explicit consent in 2024. In 2012, Alaska, California, and South Dakota required implied consent with opt-out, but Alaska changed to mandatory in 2018, followed by California and South Dakota in 2024. Missouri made participation mandatory in 2012 and 2018, but switched to implicit consent in 2024.

### Data sharing regulations

The number of jurisdictions with data sharing regulations increased from 35 (66%) in 2012 to 58 (91%) in 2024 (**Table 1, Figure 1-E**). Among all states with a regulation in immunization data sharing in 2024, some of the states, such as Alabama, Connecticut, and Florida, changed their data sharing policy from allowing everyone to access the IIS data without a permit in 2012 to requiring a data sharing agreement and protection of data’s confidentiality by 2024. States like Hawaii, Kansas, and Louisiana require that data are only used for public health or other specific purposes. Other states such as California, Indiana, and Minnesota allow data sharing only upon the request or with approval from the State Health Department.

### Temporal trends in IIS regulations

Many states and jurisdictions changed IIS regulations over time across five key policy areas (**Table 2**). Reporting mandates changed over time, with 33 states and jurisdictions (61%) showing an increased/expanded trend by mandating reporting, while only Maine (2%) showed a reduced/narrowed trend (**Figure 1-B**). Specifically, Maine transitioned from a mandate in 2000 to no mandate in 2012 and 2018, and to limited reporting requirements for certain providers and immunizations in 2024. Consent requirements also varied over time. For children, 23 states and jurisdictions (43%) moved toward more restrictive consent policies, whereas 3 (5%) relaxed their requirements (**Figure 1-C**). Similarly, for adults, 10 (19%) adopted stricter consent requirements, while 3 (5%) became less restrictive (**Figure 1-D**). Regarding data sharing policies, 19 states and jurisdictions (35%) had data sharing policies added over time, while 2 (4%) had such regulations removed (**Figure 1-E**). Unlike other policy areas, age group coverage remained largely unchanged, with 52(96%) states and jurisdictions maintaining the same policy over time and only 2 (4%) expanding coverage.

**Table 2.** Trends in changes among states and jurisdictions that had more than two data points between 2000 and 2024.

| Characteristics | No changes over time | Increased/expanded trend | Reduced/narrowed trend |
| --- | --- | --- | --- |
| Age groups | 52 (96%) | 2 (4%) | 0 (0%) |
| Provider reporting mandate to IIS or the State Health Department | 20 (37%) | 33 (61%) | 1 (2%) |
| Child's consent to participate in IIS | 28 (52%) | 23 (43%) | 3 (5%) |
| Adult consent to participate in IIS | 41 (76%) | 10 (19%) | 3 (5%) |
| Data sharing policy | 33 (61%) | 19 (35%) | 2 (4%) |

An “increased/expanded” policy trend refers to expanded population enrollment in state IIS; shifts in regulations from IIS participation with explicit consent to implicit consent to mandatory participation without consent; and the introduction of data sharing regulations.
A “reduced/narrowed” policy trend refers to narrowed population enrollment in state IIS; shifts in regulations from mandatory participation without consent to participation with implicit consent to explicit consent; and the removal of data sharing regulations.

A complete list of key characteristics of IIS regulations in each state and jurisdiction for the years 2000, 2012, 2018, and 2024 can be found in **Supplementary Data**.

## Discussion

This study describes how IIS policies have changed across U.S. states and jurisdictions between 2000 and 2024. The most consistent change over time was the expansion of provider reporting mandates, whereas other key areas, especially consent policies, remained heterogeneous and, in some cases, moved in opposing directions across jurisdictions. These temporal changes and heterogeneity should be considered when interpreting IIS data, particularly in longitudinal and cross-jurisdictional analyses.

Our findings show important progress in the scope of IIS over time. While some states limited IIS enrollment to children until 2018, all states and jurisdictions authorized coverage across the lifespan in 2024. The number of states and jurisdictions mandating provider reporting increased from 12 in 2000 to 54 in 2024. These shifts indicate broad acceptance of IIS as critical for capturing comprehensive vaccination data and supporting public health surveillance. At the same time, some variation remains across key policy areas. Jurisdictions continue to vary in which providers are required to report. Consent frameworks also remain heterogeneous, with most states adopting implicit consent with opt-out provisions, while others continue to require explicit opt-in consent, particularly for adults.

IIS provides important data for vaccine evaluation. IIS data have been shown to provide more complete vaccination records than self-reported data, which are subject to recall bias, supporting their use in epidemiologic research [12]. Despite their potential, however, IIS data have remained underutilized in research. Previous studies have found that only a small proportion of published studies use IIS data, often limited to a few jurisdictions and older data [12,13]. Variation in data sharing policies across jurisdictions and operational constraints, such as limited staffing and infrastructure, were highlighted as barriers for the broader use of IIS data for research. We found that many states introduced data-sharing policies over time, which represents important progress. When analyzing data across states and over time, however, differences in consent requirements and reporting mandates may need to be considered, as they may affect data completeness and comparability. These differences may also influence vaccine effectiveness estimates derived from IIS-linked data. For example, incomplete provider reporting or lower participation in jurisdictions with more restrictive consent requirements could result in underascertainment of vaccination records and misclassification of vaccinated individuals as unvaccinated. Previous work has shown that IIS participation policies and reporting requirements may contribute to incomplete vaccination capture and variation in IIS-based coverage estimates across states [14]. Such misclassification may bias vaccine effectiveness estimates, although the magnitude and direction of bias would likely depend on the study design and whether missing vaccination data differ systematically between cases and controls. Future research is needed to evaluate how differences in IIS regulations influence data quality, vaccine evaluation, and broader public health research applications.

This study has limitations. Data from the original sources were derived from self-reported surveys, which may be subject to misclassification or incomplete reporting. In addition, the scope and detail of survey instruments also varied across years. Despite these limitations, this research still provides the most comprehensive longitudinal look at IIS to date.

## Conclusion

Over the past two decades, legislation, regulations, and policies related to IIS have changed, with a general trend toward expanded scope and participation. At the same time, substantial heterogeneity in key IIS policies remains across states. These differences, both across jurisdictions and over time, should be considered when analyzing nationwide data, particularly in longitudinal evaluations. Strengthening collaborations across IIS programs, disease surveillance teams, governmental agencies, and academic institutions may facilitate data sharing and support the broader use of IIS data for vaccine evaluation and public health research.

## Data Availability

All data produced in the present study are available upon reasonable request to the authors

## Acknowledgment

This study was made possible by a cooperative agreement CDC-RFA-FT-23-0069 from the CDC’s Center for Forecasting and Outbreak Analytics. The funders of the study had no role in study design, data collection, data analysis, data interpretation, or writing of the report. Its contents are solely the responsibility of the authors and do not necessarily represent the official views of the funders. T.C. has received financial compensation from YouTube to help them better understand vaccination in the United States.

## Supplementary Figures

**Supplementary Figure 1.**
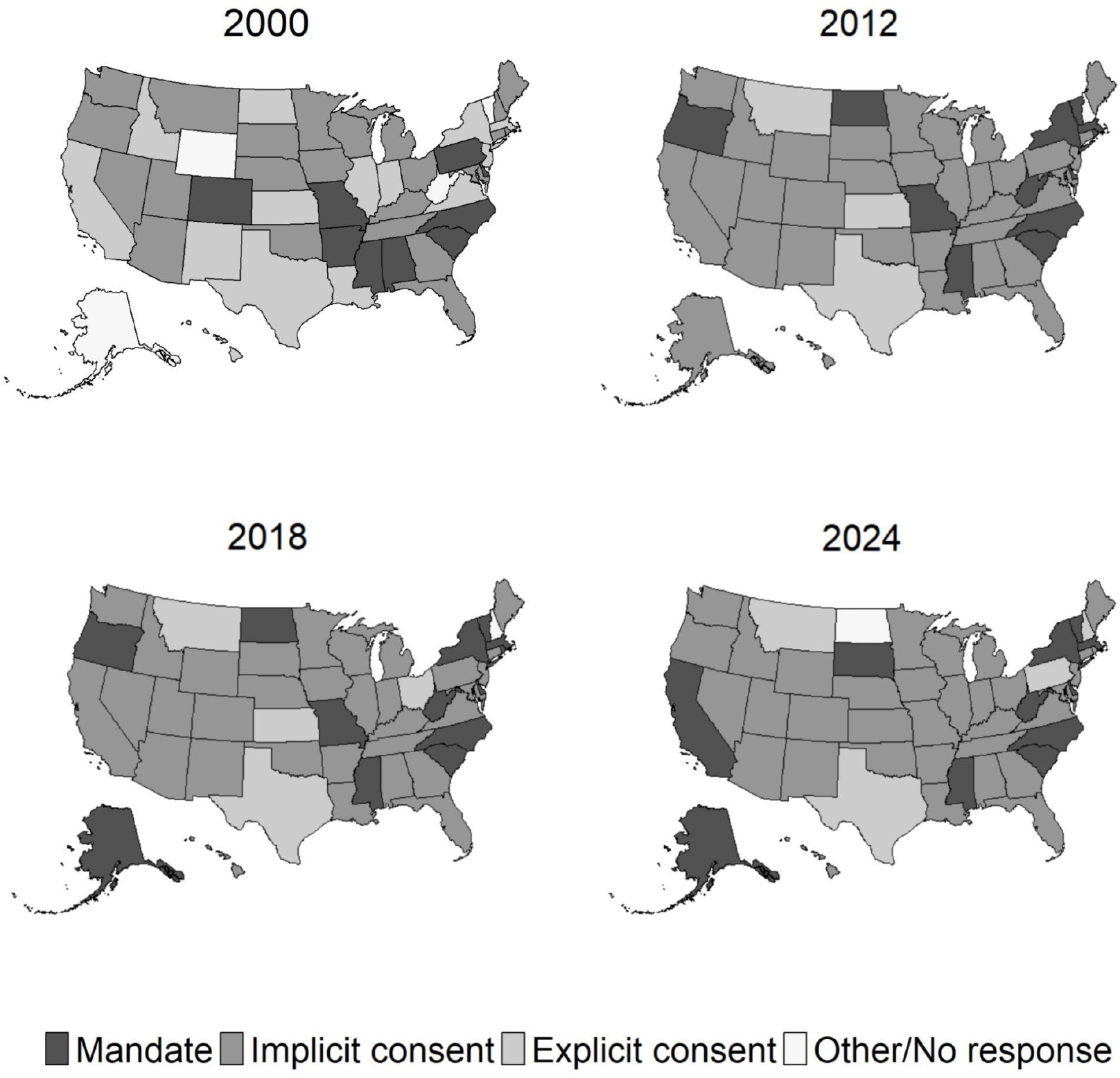
Children’s consent to participate in IIS or immunization registries in 2000, 2012, 2018, and 2024 across states. It should be noted that the immunization registry data collected in the previous study (Horlick, *et al*.) focused on state immunization registries rather than specifically on IIS. Therefore, some states may have operated their own immunization registries in 2000 without participating in the IIS network monitored by CDC. For example, New Hampshire had legislation authorizing an immunization registry in 2000, but it did not participate in IIS until later. We still included such jurisdictions in the analysis to capture temporal differences in immunization registry policies across jurisdictions. In contrast, the data collected for 2012-2024 specifically focused on IIS across jurisdictions; therefore, jurisdictions that did not participate in IIS during those years were not included (e.g., New Hampshire in 2012 and 2018).

**Supplementary Figure 2.**
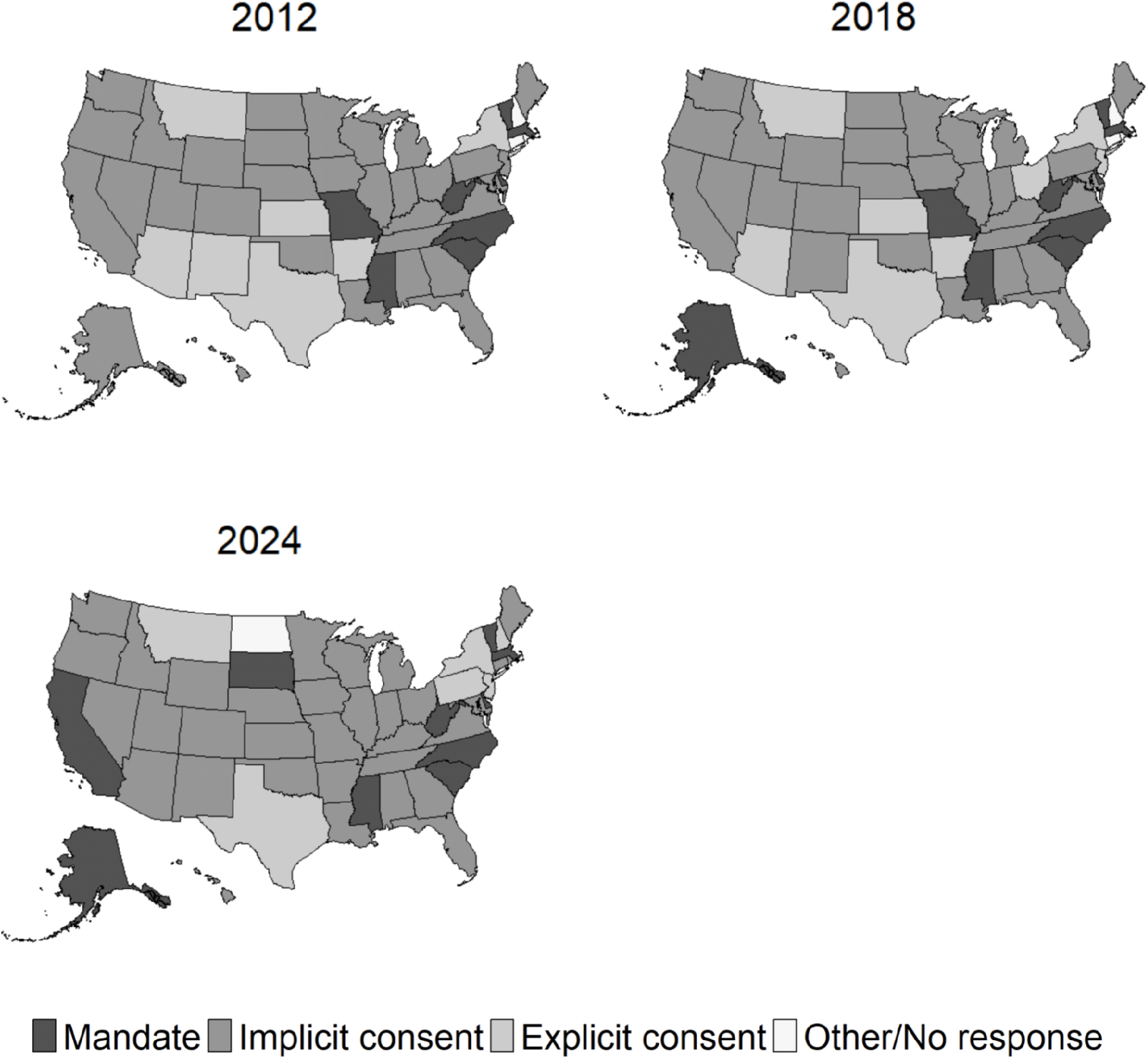
Adult consent to participate in IIS in 2012, 2018, and 2024

## References

[1] Horlick GA, Beeler SF, Linkins R. A review of state legislation related to immunization registries11SF Beeler currently works for Children’s Healthcare of Atlanta. Am J Prev Med 2001;20:208–13. 10.1016/S0749-3797(00)00309-3.

[2] Linkins RW, Feikema SM. Immunization registries: the cornerstone of childhood immunization in the 21st century. Pediatr Ann 1998;27:349–54. 10.3928/0090-4481-19980601-09.

[3] Watson WC, Saarlas KN, Hearn R, Russell R. The All Kids Count National Program: A Robert Wood Johnson Foundation Initiative to Develop Immunization Registries. Am J Prev Med 1997;13:3–6. 10.1016/S0749-3797(18)30104-1.

[4] CDC. Immunization Information Systems Resources. https://www.cdc.gov/iis/about/index.html; 2024 (accessed 30 June 2025)

[5] CDC. IIS Policy and Legislation. https://www.cdc.gov/iis/policy-legislation/index.html; 2024 (accessed 30 June 2025)

[6] Schneider KL, Bell EJ, Zhou CindyK, Yang G, Lloyd P, Clarke TC, et al. Use of Immunization Information Systems in Ascertainment of COVID-19 Vaccinations for Claims-Based Vaccine Safety and Effectiveness Studies. JAMA Netw Open 2023;6:e2313512. 10.1001/jamanetworkopen.2023.13512.

[7] Surie D, Bonnell LN, DeCuir J, Gaglani M, McNeal T, Ghamande S, et al. Comparison of mRNA vaccine effectiveness against COVID-19-associated hospitalization by vaccination source: Immunization information systems, electronic medical records, and self-report—IVY Network, February 1–August 31, 2022. Vaccine 2023;41:4249–56. 10.1016/j.vaccine.2023.05.028.

[8] Guh AY, Hadler JL. Use of the state immunization information system to assess rotavirus vaccine effectiveness in Connecticut, 2006–2008. Vaccine 2011;29:6155–8. 10.1016/j.vaccine.2011.06.066.

[9] Martin DW, Lowery NE, Brand B, Gold R, Horlick G. Immunization Information Systems: A Decade of Progress in Law and Policy. J Public Health Manag Pract 2015;21:296–303. 10.1097/PHH.0000000000000040.

[10] Kates J, Bell C, Michaud J, Williams E, Tolbert J. Tracking State Actions on Vaccine Policy and Access. KFF 2025. https://www.kff.org/covid-19/tracking-state-actions-on-vaccine-policy-and-access/ (accessed November 14, 2025).

[11] CDC. Survey of State Immunization Information System Legislation. https://www2a.cdc.gov/vaccines/iis/iissurvey/Legislation-survey.asp; 2018 (accessed 30 June 2025).

[12] Nowalk MP, D’Agostino HEA, Zimmerman RK, Saul SG, Susick M, Raviotta JM, et al. Agreement among sources of adult influenza vaccination in the age of immunization information systems. Vaccine 2021;39:6829–36. 10.1016/j.vaccine.2021.10.041.

[13] Curran EA, Bednarczyk RA, Omer SB. Evaluation of the frequency of immunization information system use for public health research. Human Vaccines & Immunotherapeutics 2013;9:1346–50. 10.4161/hv.24033. (accessed October 17, 2025).

[14] Scharf LG, Coyle R, Adeniyi K, Fath J, Harris L, Myerburg S, et al. Current Challenges and Future Possibilities for Immunization Information Systems. Academic Pediatrics 2021;21:S57–64. 10.1016/j.acap.2020.11.008.

